# A Recovery Algorithm and Pooling Designs for One-Stage Noisy Group Testing under the Probabilistic Framework

**DOI:** 10.1101/2021.03.09.21253193

**Authors:** Yining Liu, Sachin Kadyan, Itsik Pe’er

## Abstract

Group testing saves time and resources by testing each pre-assigned group instead of each individual, and one-stage group testing emerged as essential for cost-effectively controlling the current COVID-19 pandemic. Yet, the practical challenge of adjusting pooling designs based on infection rate has not been systematically addressed. In particular, there are both theoretical interests and practical motivation to analyze one-stage group testing at finite, practical problem sizes, rather than asymptotic ones, under noisy, rather than perfect tests, and when the number of positives is randomly distributed, rather than fixed.

Here, we study noisy group testing under the probabilistic framework by modeling the infection vector as a random vector with Bernoulli entries. Our main contributions include a practical one-stage group testing protocol guided by maximizing pool entropy and a maximum-likelihood recovery algorithm under the probabilistic framework. Our findings highlight the implications of introducing randomness to the infection vectors – we find that the combinatorial structure of the pooling designs plays a less important role than the parameters such as pool size and redundancy.

## 1 Introduction

Group testing is a procedure to find positives in a cohort by applying tests for the presence of any positives to cohort subsets (groups), instead of testing each individual separately. When the fraction of positives among the samples is low, implementing group testing saves time and resources. Group testing has applications in genetics [21], drug screening [13] communications [25] and epidemiology [23].

One-stage group testing [6] addresses the scenario of groups being set in advance, independently of test results - a common practical requirement, e.g. due to testing speed constraints. Existing algorithms on infection vector recovery for noisy group testing under the combinatorial prior (the number of positives among samples is fixed) include LP relaxation [16], belief propagation [20], Markov Chain Monte Carlo (MCMC) [9], Noisy Combinatorial Orthogonal Matching Pursuit (NCOMP) [4], separate decoding [18] and Definite positives (DD) [19]. However, group testing under a fully probabilistic framework has not been extensively studied. In particular, to the best of our knowledge, there has been a lack of work on non-asymptotic results on noisy group testing in the realistic scenario where number of positives in not known in advance, as in [2, 3], but rather is a random variable.

As part of the global efforts to control the COVID-19 pandemic, there has been an emerging body of work on implementing one-stage group testing for COVID-19 [10, 11, 22–24]. However, testing scenarios vary significantly due to fluctuating infection rates across time and geography often caused by emerging variants, as well as different testing objectives, such as screening of health care workers versus large scale monitoring of the community. Group testing protocols should thus be adjusted according to the infection rate among the tested samples, measurement error rates, and the recovery error tolerance level [5].

As a result, systematically studying one-stage noisy group testing with a random number of positives is of both theoretical interests and practical importance. In this paper, we study noisy group testing under the probabilistic framework in order to address the practical challenges posted by group testing implementation for COVID-19.

The rest of the paper is organized as follows. We begin with an introduction of the noisy group testing under the probabilistic framework in Section 2, including a group testing protocol and a novel recovery algorithm under the probabilistic framework. The performance of the recovery algorithm and the pooling designs is in Section 3. Finally, we conclude with a discussion on future work in Section 4.

## 2 Methods

### 2.1 Noisy Group Testing under the Probabilistic Framework

We assume a tested cohort of *n* individuals, with some infection rate *f* among the population the cohort is sampled for. Note, that the actual number of true positives is not known in advance, as combinatorial priors unrealistically assume [2, 3]. Also, this is the ground truth rate of infected individuals, as opposed to the observed positivity rate. Specifically:

#### Definition 1.

*An infection vector X is a random vector with n i*.*i*.*d. Bernoulli*(*f*) *entries*.

We aim to design a protocol to test *n* samples with *t < n* tests. We arrange the pool assignments into a pooling matrix *M* ∈ {0, 1} ^*t×n*^ such that *M*_*ij*_ = 1 if and only if individual *j* is included in pool *i*. Notice that each row *i* sum of *M* correspond to a pool size *s*_*i*_, and each column *j* sum corresponds to the number of pools that a sample participates, which we define as redundancy *r*_*j*_.

Following [1] and [8], we focus on the following four classes of pooling designs that are of practical interests (see Fig. 1):

**Fig. 1.**
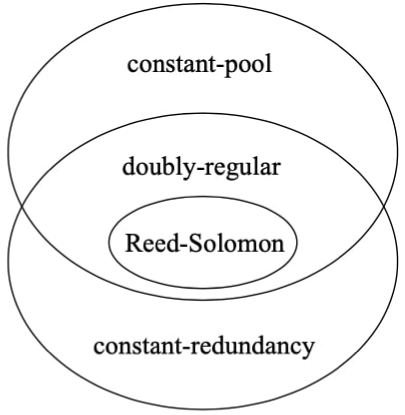
Four classes of pooling designs.

1. (constant-pool design) each test combines a fixed number *s*_*i*_ = *s* of samples, where *s* samples are chosen uniformly at random among the *n* samples;
2. (constant-redundancy design) each individual participates in a fixed number *r*_*j*_ = *r* of tests, where *r* tests are chosen uniformly at random among the *t* tests;
3. (doubly-regular design) draw a design uniformly at random from the designs that are both constant-pool and constant-redundancy;
4. (Reed-Solomon design) the pool assignment for each individual is obtained as a concatenation of Reed-Solomon error correcting code; see [8] for more details on the explicit construction.

Under the noiseless setting, a test result is negative if and only if all samples in the pool are negative. In practice, test results might suffer from measurement errors. Suppose the test we use for each pool has a false negative rate of *f*_0_ and a false positive rate of *f*_1_, i.e. consider the asymmetric noisy channel (where *f*_0_ and *f*_1_ can be distinct) that combines the additive model and the dilution model in [2]. See Fig. 2 for an example of a doubly-regular pooling matrix under the probabilistic framework.

**Fig. 2.**
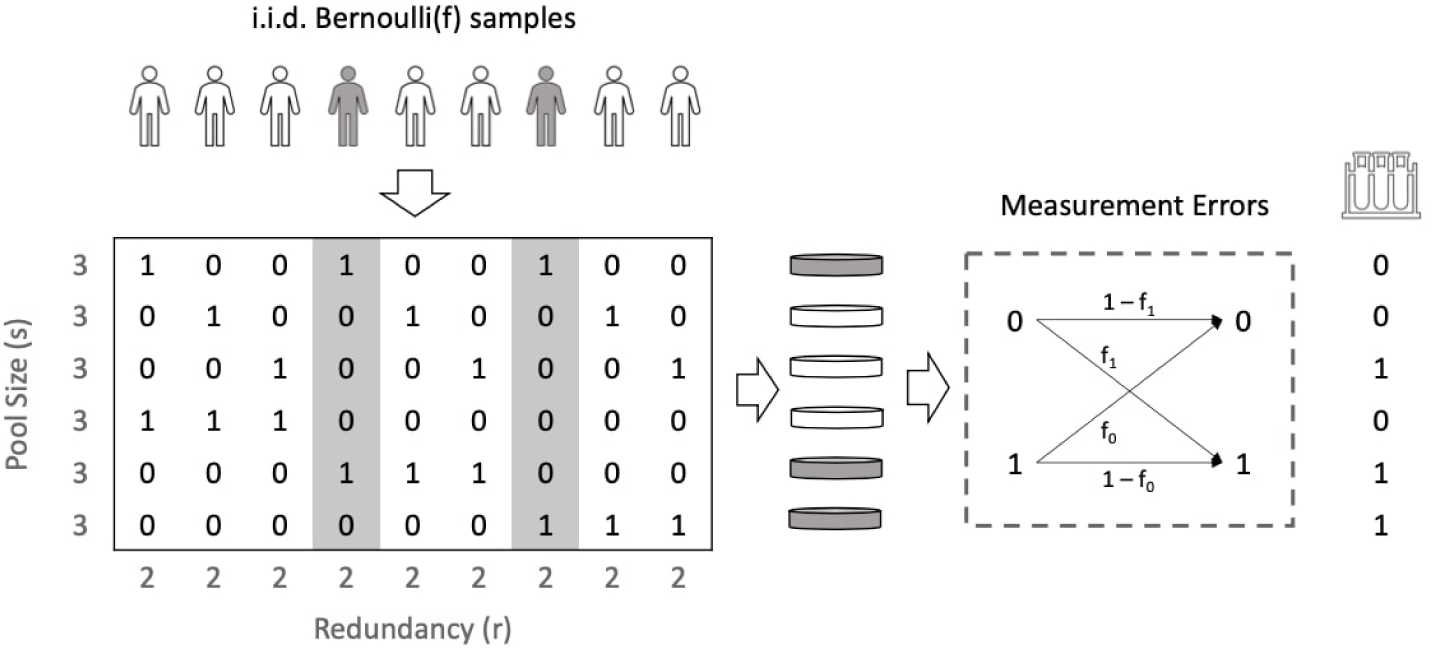
Noisy group testing under the probabilistic framework. The pooling matrix is a doubly-regular design with *s* = 3 and *r* = 2. A pool is negative if and only if there is no positive sample participates in the pool. Each pool is then passed to an asymmetric noisy channel to model measurement errors.

### 2.2 A Group Testing Protocol

Let *Y*_*i*_ be the indicator that the test result for the pool i is negative; then

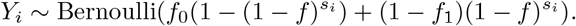

Extending the idea in [22] to the noisy setting, a pool design should maximize the entropy of *Y*_*i*_; hence the optimal pool size is

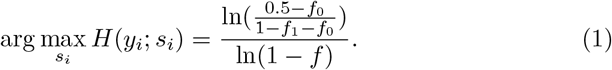

Infection rates therefore dramatically affect the theoretical optimal pool size, while measurement errors slightly offsets such optima (see Fig. 3).

**Fig. 3.**
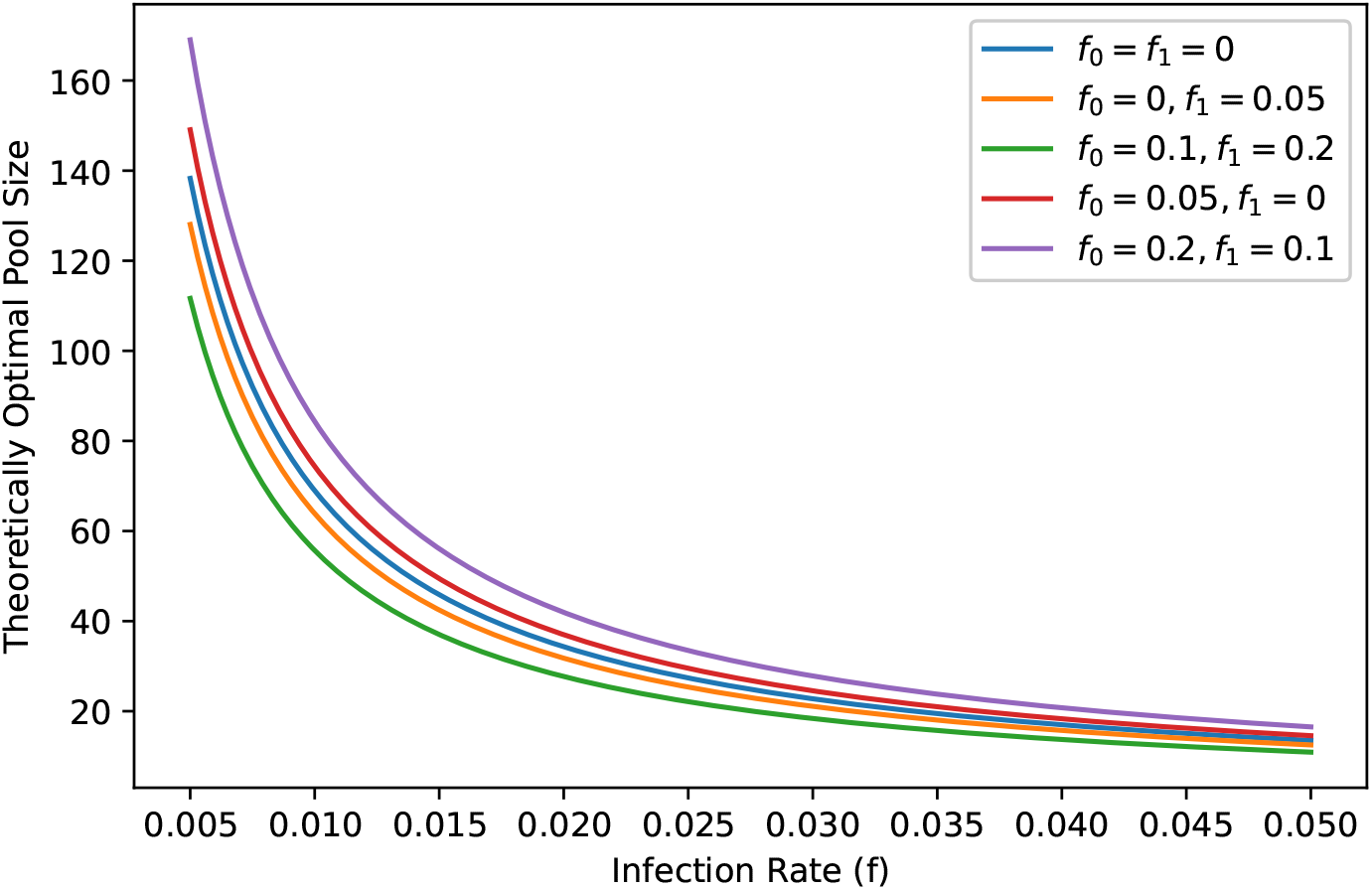
Theoretically optimal pool size as a function of infection rate; false positives (resp. negatives) slightly reduce (increase) optimal pool size.

We will show in Section 3 that given a fixed set of parameters *r, s, t*, the four classes of designs (and that of each member of a given class) perform comparably under this probabilistic framework. Hence, practitioners can safely choose a design class with practical advantages and then optimize the parameters *r, s, t*.

We thus propose the following protocol for implementing one-stage group testing under a given infection rate:

1. From the potential pool sizes (subject to practical constraints), choose *s* to be closest to the theoretical optimal pool size, per Equation 1.
2. Simulate the performance curve using the recovery algorithm in Section 2.3.
3. Choose the number of pools *t* based on the error tolerance level.
4. Perform one-stage group testing with the same recovery algorithm in step 2.

### 2.3 Recovery Algorithm

We now introduce an algorithm to recover the infection vector for noisy group testing under the probabilistic framework.

Assume we are given a pooling design *M* with *t* pools; the pool results is

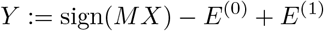

where *E*^(0)^, *E*^(1)^ are false-negatives vector and false-positives vector respectively, i.e. for *i* = 1, …, *t*,

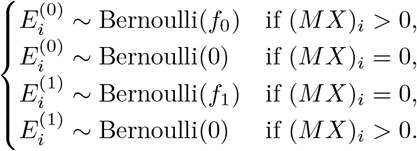

A recovery algorithm is given a pool result *y* ∈ {0, 1}^*t*^ and outputs (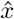 *ê*^(0)^, *ê*^(1)^), an estimate for (*X, E*^(0)^, *E*^(1)^). Our recovery algorithm (**Algorithm 1**) is given by solving the following integer linear program:

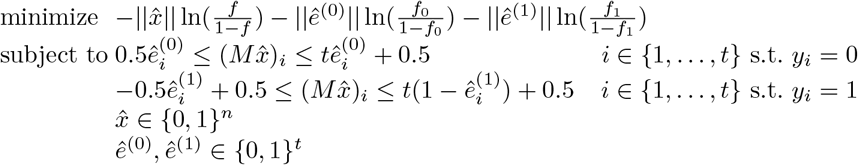

Here, for a given binary vector *x* ∈ {0, 1}^*n*^, we use ||*x*|| to denote the Hamming weight of *x*, i.e. 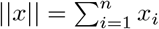

We now analyze the correctness and optimality of Algorithm 1.

#### Theorem 1.

*Algorithm 1 returns a realizable output*.

*Proof*. In order for an output (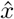, *ê*^(0)^, *ê*^(1)^) to be realizable, each row of the equation 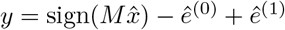 must fall into one of the following cases:

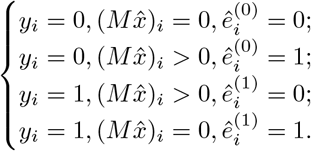

Notice that for *y*_*i*_ = 0, we have

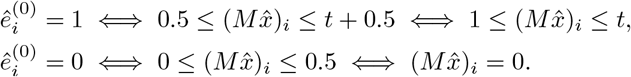

Similarly, for *y*_*i*_ = 1, we have

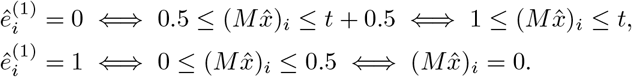

Therefore, the constraints in the integer linear program guarantees the output is realizable.

#### Theorem 2.

*Algorithm 1 returns a maximum likelihood estimate (MLE), i*.*e. an output (*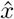, *ê*^(0)^, *ê*^(1)^*) maximizes* Pr(*X* = *x, E*^(0)^ = *e*^(0)^, *E*^(1)^ = *e*^(1)^).

*Proof*. Let *n*_0_ be the number of pools with no positive sample and *n*_1_ be the number of pools containing at least one positive sample. The log-likelihood is

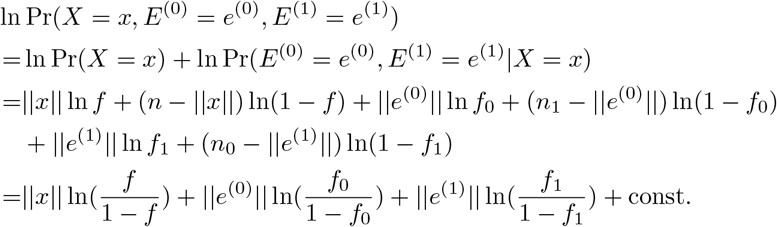

Therefore, the objective function in the integer linear program maximizes the likelihood.

Open source code implementing all our methods is available at [15].

## 3 Results

For biological experiments, sample sizes *n* = 96, 384, 1536 are of particular interests because the experiments tend to be conducted on 96-well plates, 384-well plates, or 1536-well plates [8]. The savings in resources offered by pooled design compared to individual testing stops being worth the increased complexity of the experimental paradigm when infection rates are high. For example, under an infection rate *f* = 0.1, using a constant-pool design with optimal pool size *s* = 7, even a 2-fold saving, with *T* = *n/*2 = 192 pools to recover *n* = 384 samples, the average accuracy over 1000 trials is unacceptably low at 96.5%. As a result, we believe our group testing protocol should not be applied when the infection rate is over 0.1, and limit our analysis to infection rate up to 0.025 in this section.

### 3.1 Performance of the Recovery Algorithm

In order to test the performance of the recovery algorithm, we used the pooling design *M* ∈ {0, 1} ^48*×*384^ (testing 384 individuals in 48 pools) in [23].

We simulated 1000 infection vectors *x*∈ {0, 1} ^384^ under the population infection rate *f* = 0.02 and evaluated recovery under different rates of false positives and false negatives. We observe error rate to only slightly detract from the accuracy of noiseless measurements (see Fig. 4).

**Fig. 4.**
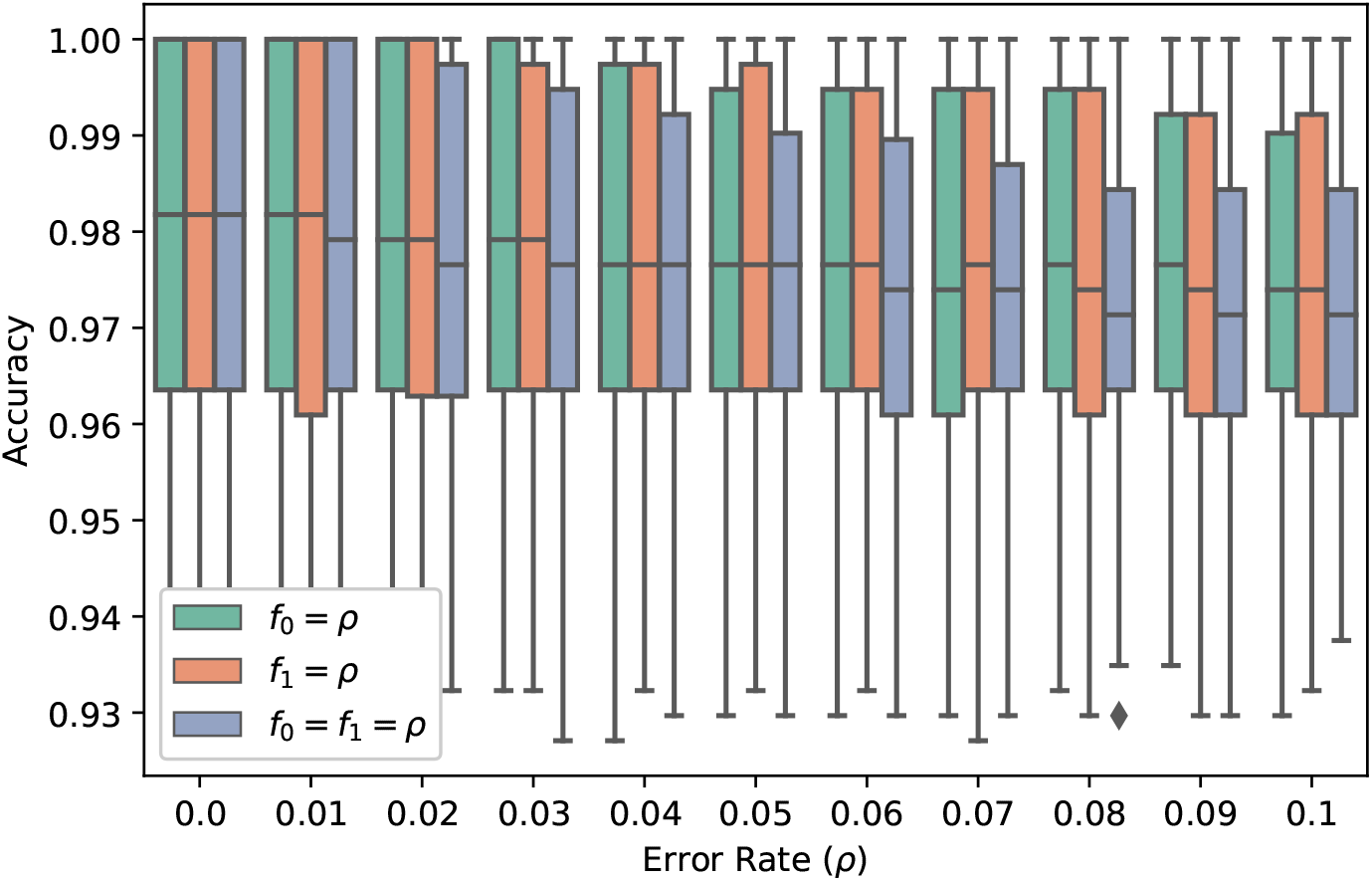
Robustness against measurements errors.

### 3.2 Pooling Matrices

#### Optimal Pool Size

In order to test the theoretically optimal pool size given in Equation 1, we simulated 1000 infection vectors under various infection rates and evaluated the performance of constant-pool matrix with *t* = 48 and *s* ∈ {8, 16, 24, …, 80}. The performance of the pooling matrices is strongly correlated with the entropy of each pool, which supports the guidance of maximizing pool entropy in one-stage group testing designs (see Fig. 5).

**Fig. 5.**
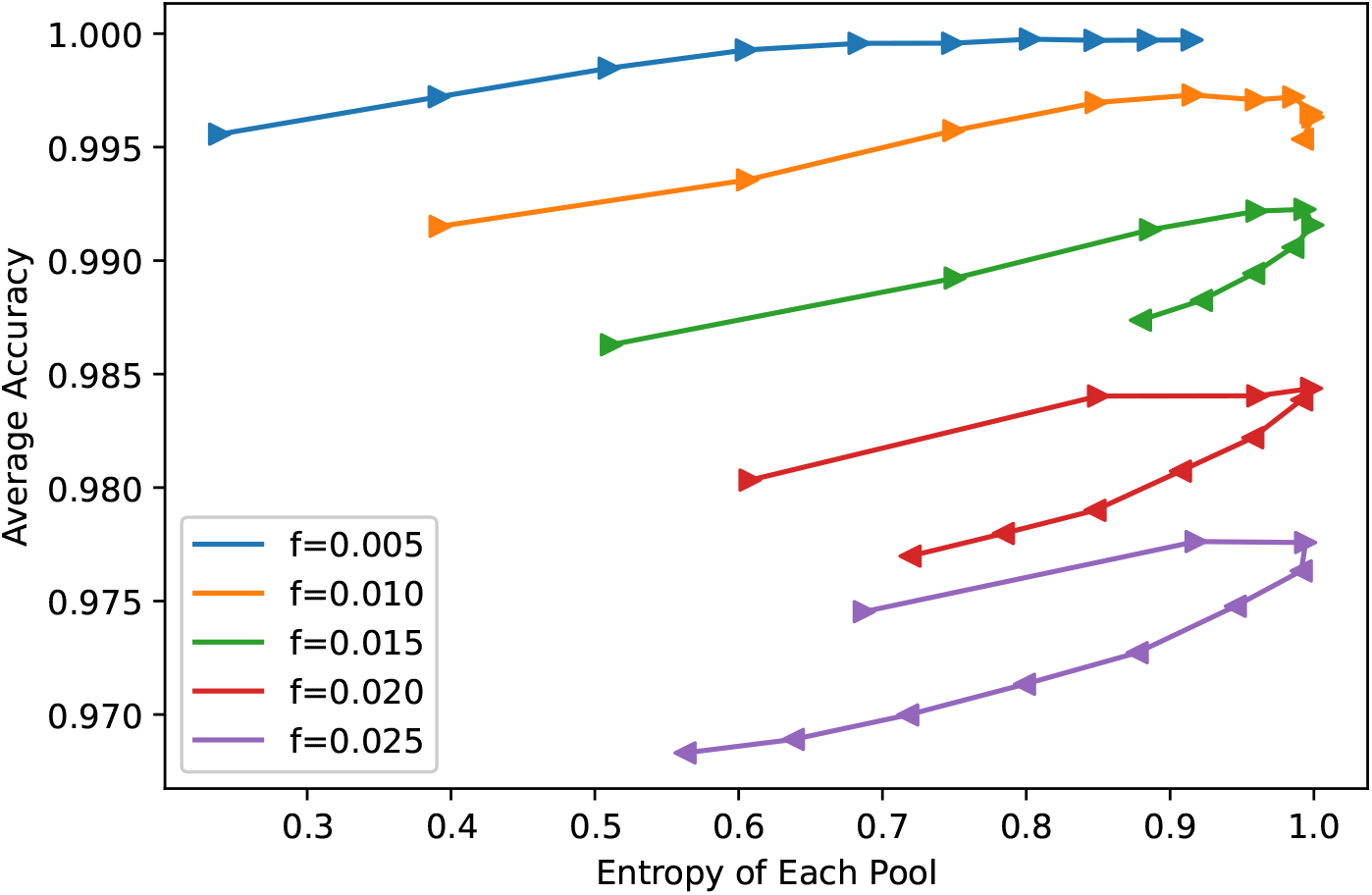
Entropy of each pool against the performance of constant-pool designs with pool size *s* ∈ {8, 16, …, 80}and *t* = 48 pools; infection vectors are simulated with 384 Bernoulli(*f*) entries. The direction of the triangular markers indicates increasing *s*.

In order to test the robustness of the optimal pool size *s* with respect to varying the number of pools *t*, we tested the performance of constant-pool designs with *s* ∈ {8, 16, 24, …, 80} based on 1000 simulated infection vectors under *f* = 0.02. The optimal pool size (*s* = 32) achieves nearly maximal recovery accuracy for all *t* (see Fig. 6).

**Fig. 6.**
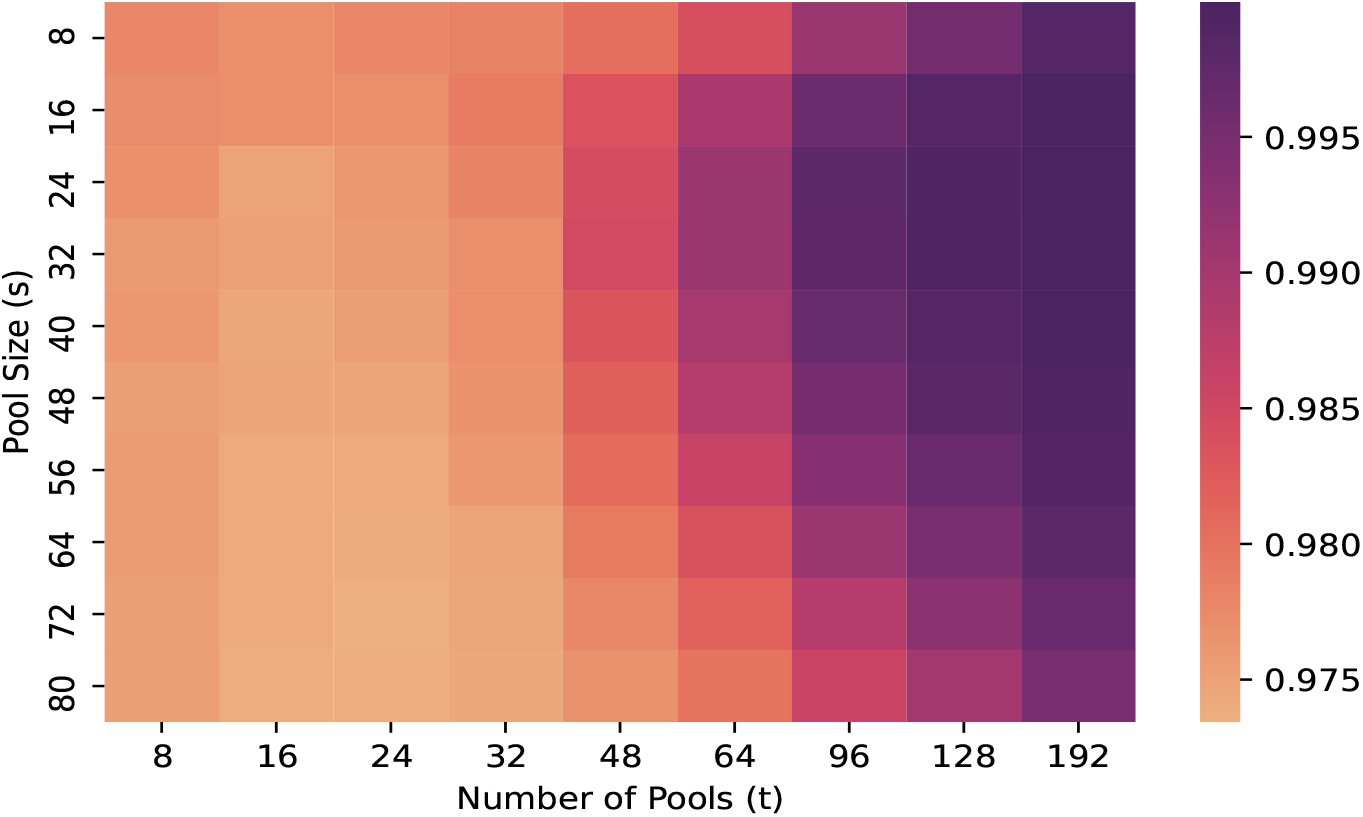
Average recovery accuracy of designs with the pool size *s* ∈ {8, 16, 24, …, 80} and varying the number of pools under the infection rate *f* = 0.02; the pool size *s* = 32 achieves the maximum pool entropy.

#### Explicit Construction

Combinatorial structure of pooling matrices has been studied under the combinatorial framework [7]. Recent development of explicit pooling matrices construction for noisy group testing is based on error correcting codes [1]. Pooling design based on Reed-Solomon code was introduced in [14] and shown to be optimal under a more restricted probabilistic setting (assuming a random set of positives of a fixed size) [12].

Authors in [23] use a design based on Reed-Solomon error correcting code; the matrix assigns each individual in 6 pools such that each pool has 48 individuals. We compare the performance of the Reed-Solomon design, a randomly drawn doubly-regular design (with *r* = 6 and *s* = 48), a randomly drawn constant-redundancy design (with *r* = 6 and varying pool sizes), and a randomly drawn constant-pool design (with *s* = 48 and varying redundancies).

We found that under the probabilistic framework, the performance of the three random designs is comparable to that of the Reed-Solomon design in terms of both the average and the standard deviation of the recovery accuracy (see Fig. 7). For each of the three random designs, we computed the distribution of the inner products of column vectors (which are derived from code words) of 1000 randomly-drawn matrices. The performance guarantees of Reed-Solomon design under the combinatorial setting follows from the minimum pairwise distance of the code words [1]. The authors in [17] studied explicit pooling matrices and showed the dependency of the error probability on both the minimum and the average distance under the assumption of a fixed number of positives. In the same spirit, we found that maximizing the number of pairs of column vectors with zero inner products could lead to performance guarantees under the probabilistic framework (see Fig. 8).

**Fig. 7.**
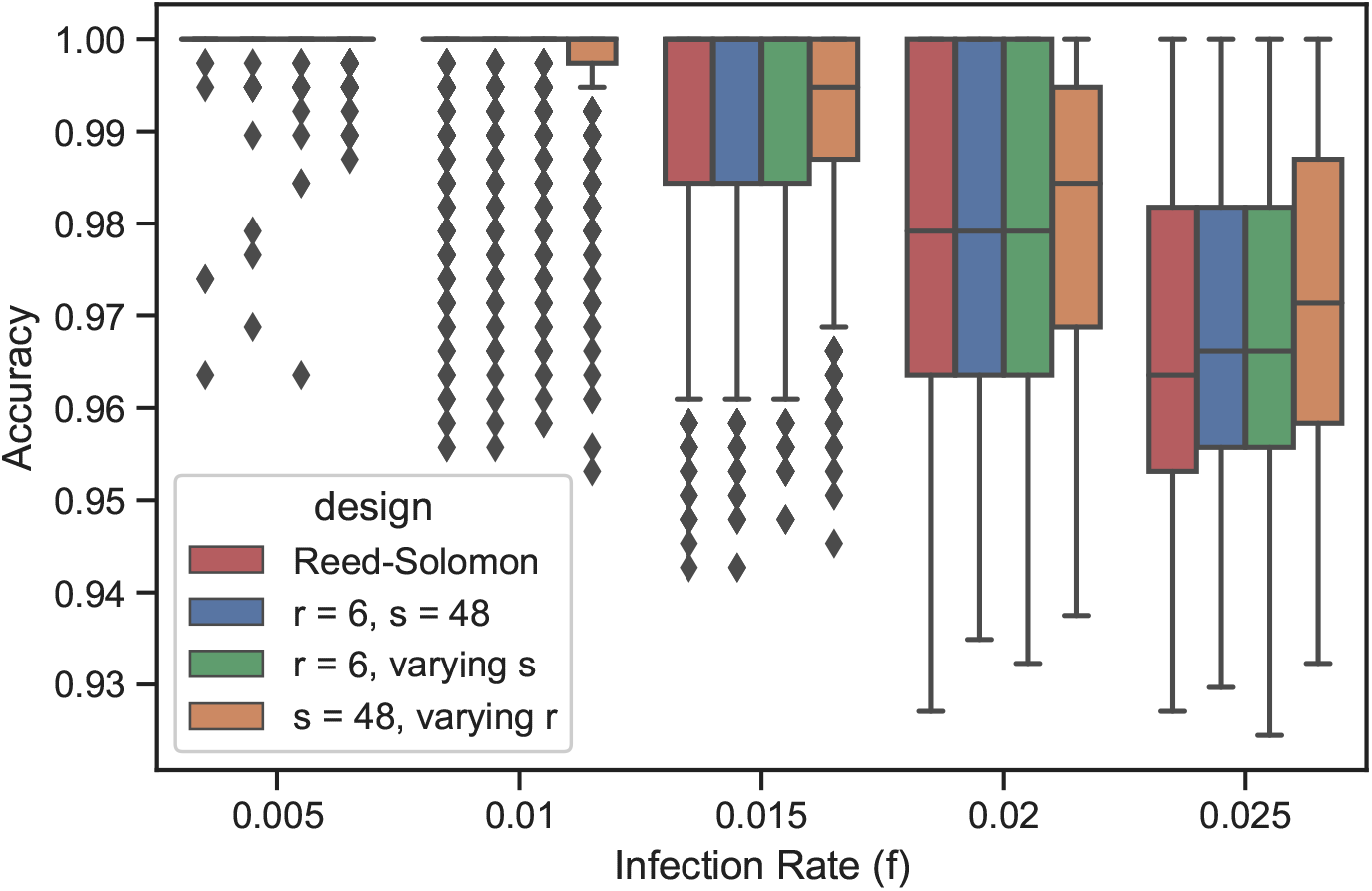
Performance of the Reed-Solomon (red), the doubly-regular *r* = 6, *s* = 48 (blue), the constant redundancy *r* = 6 (green), and the constant pool *s* = 48 (orange) designs under simulated data with *n* = 384 individuals, *t* = 48 pools. The box shows the four quartiles, the whiskers show the rest of the distribution, and the points show the outliers.

**Fig. 8.**
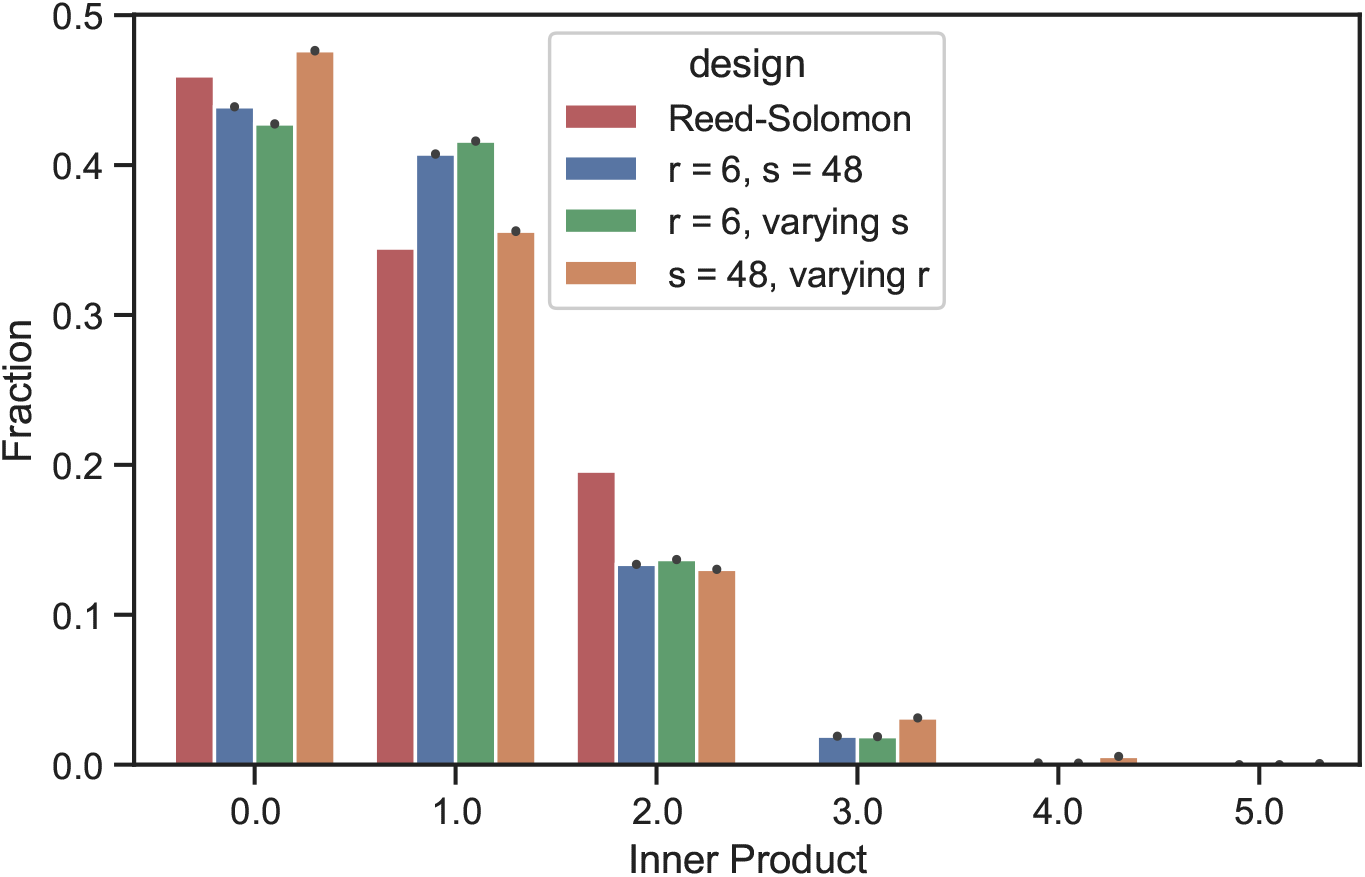
Distributions of the inner products of column vector. Error bars are computed based on 1000 matrices.

As a first step towards understanding the role of combinatorial structures of the pooling matrices under the probabilistic framework, we randomly drew 1000 constant-pool designs with *s* = 48 to test *n* = 384 individuals with *t* = 48 pools, and computed the average accuracy on two sets of 1000 simulated infection vectors under the infection rate *f* = 0.02. We observe low correlation (≈ 0.0884; *p* ≈ 0.00516) between the performance of designs on the two sets of vectors, which suggests that under the probabilistic framework, the combinatorial structure of the pooling matrices plays a less significant role than the parameters *s, r, t*.

## 4 Discussion

We presented a framework that addresses several practical issues, including randomness of the infection vector and noisy measurements. Our findings suggest that in practice, the combinatorial structure of pooling design matrices plays a lesser role compared to parameters such as redundancy, pool sizes, and the number of pools. Furthermore, we provide a protocol and an implementation for practitioners to choose the parameters in order to design pooling matrices.

From a theoretical perspective, there are several promising future directions for this work. The ILP problem presented in Section 2.3 can be viewed as a generalization of the smallest satisfying set (SSS) problem [1], which is NP-hard. One direction of future work is to design polynomial-time approximation recovery algorithm under the probabilistic framework.

In addition, non-asymptotic theoretical understanding of one-stage group testing under the probabilistic framework is still limited. An interesting future research direction is to explore theoretical justifications on the effect of the number of pairs of column vectors with zero inner product on the performance, as observed in Section 3.

From a practical perspective, while no significant difference in performance among the four classes of designs is observed in the simulations performed in this study, future research should be conducted on further investigating the significance of the combinatorial structure of the pooling matrices under other scenarios such that parameter values with a higher range of recovery accuracy and sample sizes other than *n* = 384 that are germane to biological experiments.

As the COVID-19 pandemic continue to ravage large populations of resource limited nations, cost effective testing with simple designs is an absolute necessity for sustaining daily life in a safe manner.

## Data Availability

Open source code implementing all our methods is available at https://github.com/imyiningliu/group-testing-probabilistic-framework

## Notes

### Competing Interest Statement

The authors have declared no competing interest.

### Funding Statement

No external funding was used

